# The landscape of lung bronchoalveolar immune cells in COVID-19 revealed by single-cell RNA sequencing

**DOI:** 10.1101/2020.02.23.20026690

**Authors:** Minfeng Liao, Yang Liu, Jin Yuan, Yanling Wen, Gang Xu, Juanjuan Zhao, Lin Chen, Jinxiu Li, Xin Wang, Fuxiang Wang, Lei Liu, Shuye Zhang, Zheng Zhang

## Abstract

The novel coronavirus SARS-CoV-2, etiological agent of recently named Coronavirus infected disease (COVID-19) by WHO, has caused more than 2, 000 deaths worldwide since its emergency in Wuhan City, Hubei province, China, in December, 2019. The symptoms of COVID-19 varied from modest, mild to acute respiratory distress syndrome (ARDS), and the latter of which is generally associated with deregulated immune cytokine production; however, we currently know little as to the interplay between the extent of clinical symptoms and the compositions of lung immune microenvironment. Here, we comprehensively characterized the lung immune microenvironment with the bronchoalveolar lavage fluid (BALF) from 3 severe and 3 mild COVID-19 patients and 8 previously reported healthy lung controls through single-cell RNA sequence (scRNA-seq) combined with TCR-seq. Our data shows that monocyte-derived FCN1^+^ macrophages, whereas notFABP4^+^ alveolar macrophages that represent a predominant macrophage subset in BALF from patients with mild diseases, overwhelm in the severely damaged lungs from patients with ARDS. These cells are highly inflammatory and enormous chemokine producers implicated in cytokine storm. Furthermore, the formation of tissue resident, highly expanded clonal CD8^+^ T cells in the lung microenvironment of mild symptom patients suggests a robust adaptive immune response connected to a better control of COVID-19. This study first reported the cellular atlas of lung bronchoalveolar immune microenvironment in COVID-19 patients at the single-cell resolution, and unveiled the potential immune mechanisms underlying disease progression and protection in COVID-19.

**Highlights:**

1. Immune microenvironment of SARS-CoV-2-infected lungs revealed by scRNA/TCR seq
2. Increased inflammatory FCN1+ macrophages are replacing FABP4+ macrophages in the BALF from severe COVID-19 patients
3. Highly expanded and functional competent tissue resident clonal CD8+ T cells in mild COVID-19 patients

## Introduction

Since December, 2019, a disease outbreak caused by a novel coronavirus (now given the name SARS-CoV-2) started in Wuhan, and has quickly spread in China and subsequently inmany other countries [1]. The WHO has declared the new coronavirus (CoV) infection as a global public health emergency and named the coronavirus infected disease-19 (COVID-19). The genome analysis showed that the SARS-CoV-2 is a SARS-CoV like β-lineage coronavirus [2] and likely originates from bats [3]. SARS-CoV-2 uses human ACE2 protein as their receptors [4], which explains its similarly high transmissibility. The clinical data indicated that the COVID-19 varied from asymptomatic to acute respiratory distress syndrome (ARDS), similar as SARS-CoV infection [5-7]. Although researchers are racing against time to develop vaccines and test anti-viral drugs in clinical trials [8], there is no effective prophylactic and clinical treatment for COVID-19 yet.

Generally, the COVID-19 is less severe and less fatal than the SARS, however, some patients, especially aged populations with co-morbidities are prone to develop more severe symptoms and require emergent medical interventions [9]. It is not completely understood why some patients develop severe but others have mild or even asymptomatic diseases by the same SARS-CoV-2 infections. The immunopathogenesis of hCoVs-induced respiratory distress syndrome may involve deranged interferon production, hyper-inflammatory response and cytokine storms, inefficient or delayed induction of neutralizing antibody and specific T cell responses [10, 11]. However, due to biosafety and ethics issues and technical limitations, most of the current knowledge was generated from animal model studies, and not directly from human subjects. More investigations using patient samples are needed to explore the relevant protective or pathogenic mechanisms in clinical settings.

Here, we applied the emerging single-cell RNA sequence (scRNA-seq) and single-cell TCR-seq to comprehensively characterize the lung bronchoalveolar lavage fluid (BALF) cells from 6 of COVID-19 patients, including 3 severe and 3 mild cases. Our study depicts a high-resolution transcriptome atlas of lung resident immune subsets in response to SARS-CoV-2 infections. It reveals that compared to the lung immune microenvironment of mild symptom patients, monocyte-derived FCN1^+^ macrophages replacing the FABP4^+^ alveolar macrophages predominate macrophage lineage compostions in the severely damaged lung, which are highly inflammatory and potent chemokine producers. Furthermore, the lung resident highly expanded clonal CD8^+^ T cells formed in the mildly infected patients support the notion that a rapid and robust adaptive immune response is potentially critical for controlling COVID-19.

## Results

### A glimpse of lung immune microenvironment in the COVID-19 patients

To characterize the immune microenvironment of the SARS-CoV-2-infected lung, we performed scRNA-seq analysis of single cells in the lung BALF (37, 820 cells) using the 10X Genomics platform, from 3 of recovered mild cases and 3 of severe cases (Figure 1A, Table 1). We also collected public available scRNA-seq data (43, 627 cells) of 8 normal lungs as control [12]. This dataset passed stringent high-quality filtering (Figure S1), yielding a mean of 188K reads/cell with median gene and unique molecular identifier (UMI) counts of 2, 070 and 6, 852, respectively (Table S1).

**Table 1.** Clinical data of the enrolled subjects.

|  | T141 | T142 | T143 | T144 | T145 | T146 |
| --- | --- | --- | --- | --- | --- | --- |
| Severity | mild | mild | severe | mild | severe | severe |
| Age | 36 | 37 | 66 | 35 | 62 | 63 |
| Gender | male | female | male | male | male | male |
| Wuhan exposure history | yes | yes | yes | yes | yes | yes |
| Symptom onset day | Jan 9, 2020 | Jan 11, 2020 | Jan 3, 2020 | Jan 9, 2020 | Jan 11, 2020 | Jan 8, 2020 |
| First symptom | Cough | Fever/Cough | Fever | Cough | Fever/Cough | Fever/Cough |
| Hospitalization date | Jan 16,<br>2020 | Jan 16, 2020 | Jan 11, 2020 | Jan 15,<br>2020 | Jan 18, 2020 | Jan 15, 2020 |
| Chronic basic disease | none | none | hypertension | none | none | COPD |
| SARS-CoV-2 | + | + | + | + | + | + |
| Influenza A virus | - | - | - | - | - | - |
| Influenza B virus | - | - | - | - | - | - |
| Respiratory syncytial virus | - | - | - | - | - | - |
| Adenovirus | - | - | - | - | - | - |
| Interferon atomization | Jan 16,<br>2020 | no | Jan 14, 2020 | Jan 18,<br>2020 | Jan 16, 2020 | Jan 15, 2020 |
| Ribavirin | Jan 16,<br>2020 | no | Jan 14, 2020 | Jan 18,<br>2020 | Jan 16, 2020 | Jan 15, 2020 |
| Methylprednisolone | no | no | yes | no | no | yes |
| BALF sampling date | Jan 20,<br>2020 | Jan 20, 2020 | Jan 21, 2020 | Jan 22,<br>2020 | Jan 22, 2020 | Jan 22, 2020 |
| CT finding | Bilaeral<br>pneumonia | Bilaeral<br>pneumonia | Bilaeral<br>pneumonia | Bilaeral<br>pneumonia | Bilaeral<br>pneumonia | Bilaeral<br>pneumonia<br>&<br>Bronchietasis |

**Figure 1.**
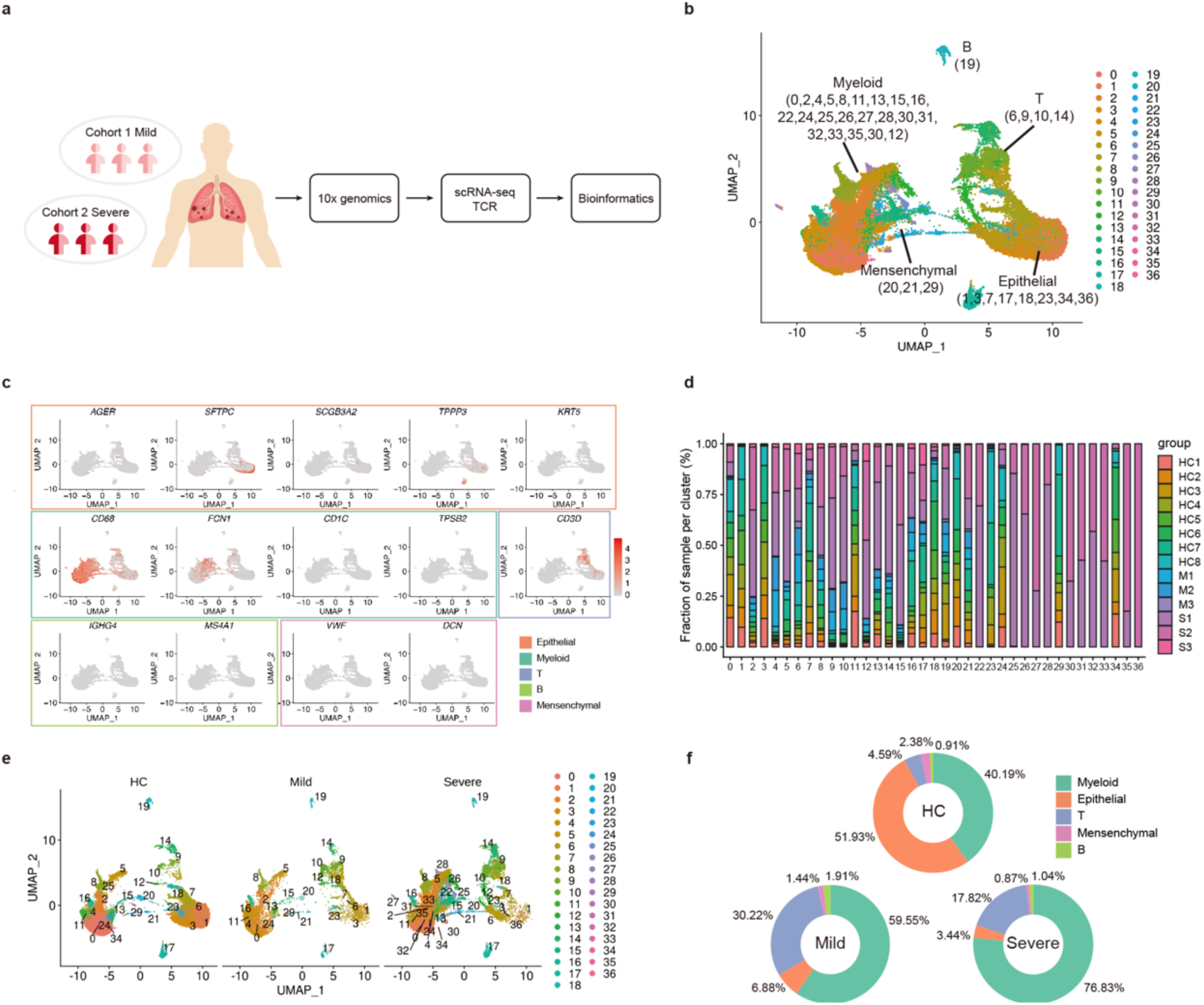
Single cell atlas of BALFs from SARS-CoV-2-infected lungs and controlled lung tissues. A. The cartoon showing the outline of study. BALFs were collected from COVID-19 patients and processed using the 10xGenomics platform for scRNA/scTCR seq. B. The UMAP presentation of single cell atlas of BALFs showing 5 major cell types and 36 clusters. Parentheses indicate the clusters of each major cell type. C. UMAP projection of canonical markers for different cell types as indicated in the legend. D. The bar plot shows the relative contributions of each cluster by individual sample. E. The UMAP comparison of the clustering distribution across control, mild and severe COVID-19 groups. F. The pie chart shows the percentages of immune and non-immune cell populations in different groups. HC1-HC8: healthy controls, M1-M3: mild cases, S1-S3: severe cases.

The clustering analysis showed 36 distinct cell clusters (C) composed of major cell subtypes including macrophages, NK & T, B and plasma cells, epithelial and endothelial cells (Figure 1B), identified by unique signature genes *CD68* (macrophages), *CD3D* and *KLRF1* (T & NK cells), *MS4A1* (B and plasma cell), *AGER, SFTPC, SCGB3A2, TPP3* and *KRT5* (epithelial cell), *VWF* (endothelial cell) and *DCN* (fibroblasts), respectively (Figure 1C and S2A). We confirmed that the data integration removed batch effects and enabled excellent reproducibility across different donors. Major cell populations and most clusters included cells from each sample (Figure 1D and S2B). We were also able to detect low levels of SARS-CoV-2 transcripts in various cell populations from severe COVID-19 patients but not mild cases and controls (Figure S2C). We assumed that these viral transcripts were likely ambient contaminations of viral loads in the BALF.

We then compared the distribution of different cellular compartments among control, mild and severe group. There were higher proportions of T and NK cells in the COVID-19 patients than those in controls, while epithelial cells in patients are fewer. As compared to mild cases, severe patients contained more macrophages but less proportion of T and NK cells (Figure 1E-1F). Together, our data showed an increased recruitment of immune cells to the lung in response to SARS-CoV-2 infection, and that the lung immune cell compartments differed between mild and severe COVID-19 patients.

### Dysregulated BALF macrophage compartments in COVID-19 patients with severe diseases

The increased lung macrophage population was present in severe COVID-19 patients. To further understand the macrophage heterogeneity, we re-clustered the macrophages and showed 22 clusters (Figure 2A). Recent studies have identified three distinct human lung macrophage subsets by *FCN1* (monocyte derived), *SPP1* (pro-fibrotic) and *FABP4* (alveolar macrophage, AM) marker expression [13]. Referring to the classification criteria and other macrophage markers (Figure S3A), we found that most macrophage clusters (except for C15 as a dendritic cell subset) in our study could also be grouped by *FCN1, SPP1* and *FABP4* expression patterns. Group 1 includes C9 and C10 (FCN1^hi^ only), Group 2 includes C0, C6, C12, C16 and C19 (FCN1^lo^SPP1^+^), Group 3 includes C7, C8 and C18 (FCN1^-^SPP1^+^), Group 4 contains C1, C2, C3, C4, C5, C11, C13, C14, C17, C20, C21, C22 (FABP4^+^) (Figure 2B).

**Figure 2.**
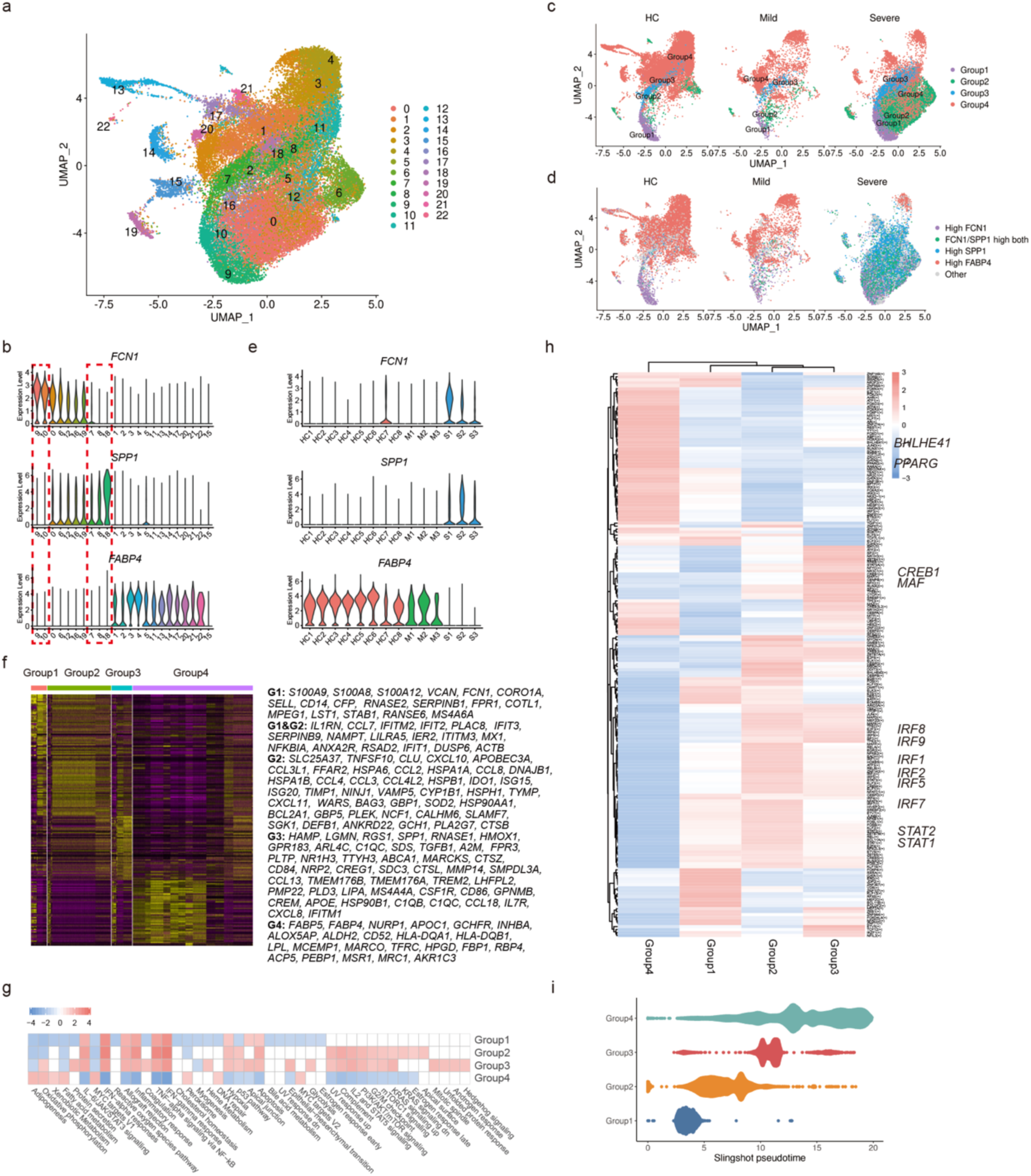
Perturbed CD68^+^ macrophage compartments in severe COVID-19. A. The UMAP presentation of the heterogenous clusters of lung macrophages B. The lung macrophages clusters were classified to 4 groups according to *FCN1, SPP1* and *FABP4* expression levels, indicated by red squares. C. The UMAP projection of four macrophage groups among controls and patients D. The UMAP projection of *FCN1, SPP1*, and *FABP4* expressed cells among controls and patients E. The violin plot shows the *FCN1, SPP1* and *FABP4* expression by lung CD68^+^ macrophages from each individual sample F. The heatmaps of hierarchically clustered top 50 differentially-expressed genes (DEGs) across 4 groups of macrophages. G. The GSEA NES heatmap of the 50 MSigDB hallmark gene sets across 4 groups of macrophages. The gene list for each group was ordered based on log2(ratio) of this group compared with all other cells. Default parameter in GSEA function of clusterProfiler was used (except nPerm = 100000). H. Average regulon activities of 4 groups of macrophages calculated with SECENIC. PySCENIC was used to infer co-expression modules, prune modules for targets with cis regulatory footprints and calculate cellular regulon enrichment matrix with default parameters. I. The differentiation trajectory of different groups of macrophages by monocle analysis. Dimensionality reduction produced by UMAP was used in lineage inference.

We found that the composition of the macrophage groups differed significantly among controls, mild versus severe COVID-19 patients (Figure 2C). This transformation was also reflected by differential expression of *FCN1, SPP1* and *FABP4* genes in the controls and patients (Figure 2D). We further confirmed that *FCN1* was preferentially expressed by individual controls and mild COVID-19 patients, while *SPP1* and *FABP4* were highly expressed by severe COVID-19 patients (Figure 2E). Together, these data showed a dysregulated balance of lung macrophage populations during the progression of severe COVID-19, manifested by substantially increased group 1 monocyte-derived macrophages (FCN1^+^), group 2 & 3 macrophages (SPP1^+^), and almost a complete loss of lung AMs (FABP4^+^).

To further understand the difference of the aforementioned 4 groups of macrophages, we conducted gene expression (GEX) analysis (Figure 2F). Group 1 expressed *S100A8, S100A9, VCAN, FCN1, CD14, SELL* associated with inflammatory monocytes. Group 2 expressed chemokines *CCL2, CCL3, CCL4, CXCL9, CXCL10, CXCL11* and ISGs *APOBEC3A, ISG15, ISG20, GBP1, ITITM3, MX1*, etc. Group 3 expressed immune regulatory genes *A2M, GPR183, CCL13* etc. and pro-fibrotic genes *TREM2, TGFB1, SPP1*. Group 4 expressed typical AM genes, such as *FABP4, FABP5, INHBA, MACRO, TFRC, MRC1*, etc. GSEA and GO analysis further revealed that group 1, 2 and 3 were more activated and inflammatory, while group 4 was featured with lipid metabolic functions (Figure 2G and S3B). Using SCENIC, we deduced the transcription factors (TFs) activity in macrophage groups (Figure 2H). Consistent with their inflammatory phenotype and functions, we found enhanced STAT1, STAT2 and multiple IFN regulatory factors (IRFs) activity among group 1, 2 and 3 macrophages. We also identified M2-like TFs, MAF and CREB1, specifically in group 3 macrophages, and AM-TFs, PPARG and BHLHE41, specifically in group 4 macrophages. Finally, we analyzed the differentiation trajectory of macrophage populations, and found a linear path starting from group 1, then 2 & 3, eventually to group 4 (Figure 2I).

Collectively, consistent with early report [13], we identified the group 1 as the monocyte-like population, group 3 as the recently reported pro-fibrotic *SPP1*^+^ population and group 4 as the lung AM. Moreover, we identified a novel intermediate macrophage population (group 2), only from the severe COVID-19 patients. Our data indicated that the monocytes are recruited from circulation (*FCN1*^+^) to the lung to fuel the inflammation during severe diseases, and some monocytes may further go through the differentiation process into the *SPP1*^+^ populations and eventually the FABP4^+^ AMs.

### Characterization of T and NK cell responses in BALF from the lungs of COVID-19 patients

NK and T lymphocytes are immune effectors against viral infection. Previous studies showed that robust and early T cell response played crucial roles in viral clearance during acute respiratory infections [14]. After exclusion of contaminated clusters, our clustering analysis of T and NK lymphocytes identified 5 major subsets (Figure 3A) based on the typical T and NK cell markers (Figure 3B). NK cells highly expressed *KLRF1, KLRC1* and *KLRD1*. CD8^+^ T cells expressed *CD8A* and *CD8B*. The CCR7^+^ cells expressing *CD4* and *IL7R* were likely CD4^+^ T cells, Treg cells were *CTLA4*^*+*^*FOXP3*^+^*IL2RA*^+^, and proliferating cells were *TYMS*^+^*MKI67*^+^.

**Figure 3.**
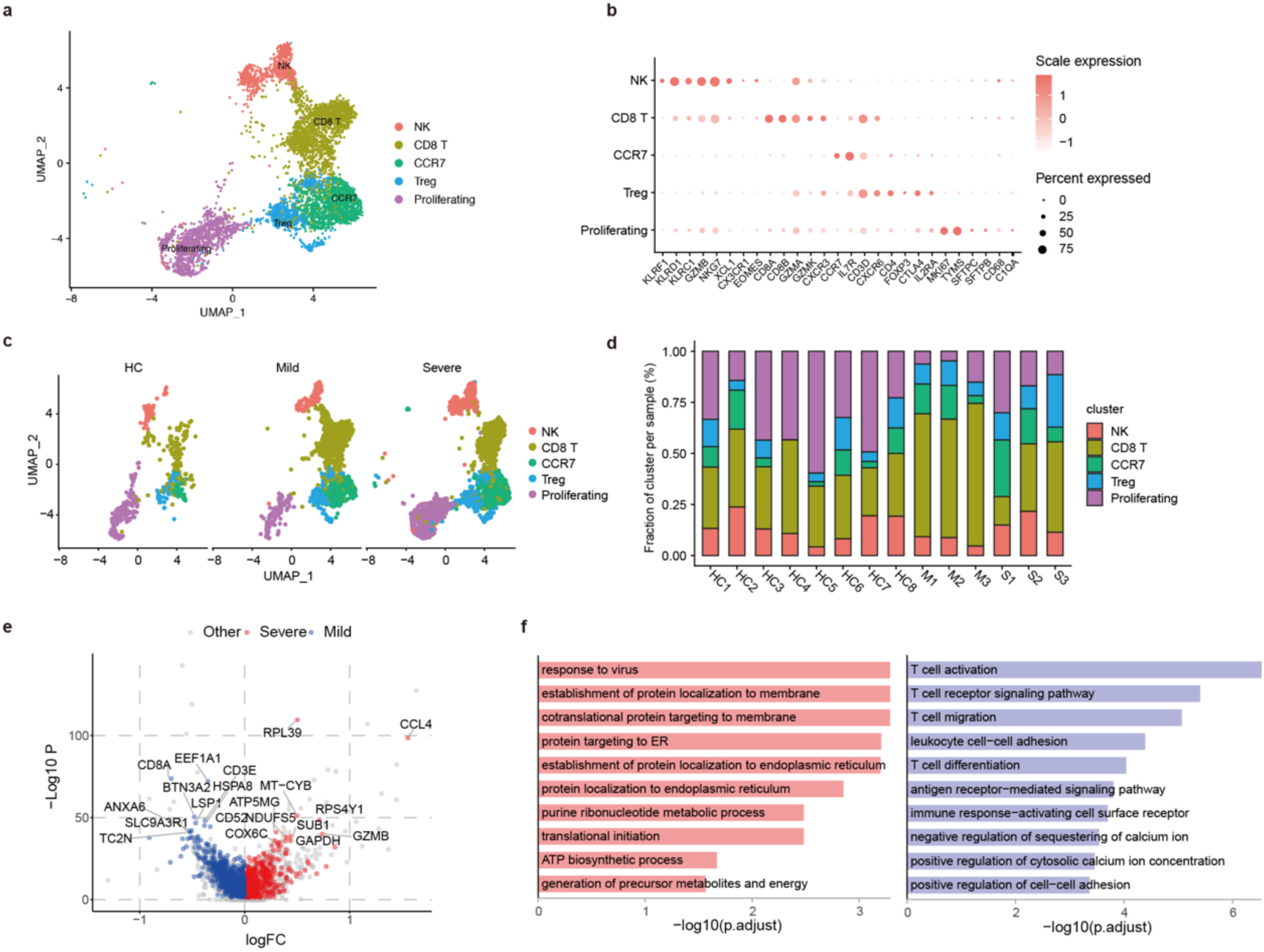
Analysis of the BALF T and NK lymphocytes in COVID-19 patients. A. The UMAP plot shows the clustering of lung T and NK cells B. The heatmap of showing the immune typing markers on different T and NK clusters C. The UMAP plots show comparison of the T and NK cell compartments across different groups D. The bar plot shows the percentages of T and NK clusters in each individual sample E. The volcano plot shows the DEGs of CD8^+^ T cells between severe vs. mild COVID-19 patients. Only genes specifically upregulated in T cells compared to macrophage cells are kept. And a gene is considered significant with adjust p < 0.05. F. The GO BP enrichment analysis of DEGs of CD8^+^ T cells between severe vs. mild COVID-19 patients with adjust p < 0.05 upregulated in severe (left) and mild (right).

Comparing few numbers of NK and T lymphocytes in controls, the proportions of NK and T lymphocytes in the lung was largely increased in COVID-19 patients (Figure 1F and 3C). Among different subsets, we observed lower proportion of CD8^+^ T cells, and increased proportion of proliferating cells in severe patients than the mild patients (Figure 3C and 3D). GEX analysis showed that CD8^+^ T cells expressed high levels of effector molecules, including *GZMA, GZMK, FASLG, CCL5*, etc., and resident T cell markers *ITGA1, CXCR6, JAML*, etc., likely played crucial roles in immune defense (Figure S4).

The higher number of lung CD8^+^ T cells in mild patients may indicate a role of CD8^+^ T cells in the clearance of the virus. In contrast, the higher level of T cell proliferation in severe disease group is intriguing. We assumed that it might be related to bystander activation induced by hyper inflammation, or a delayed response to the infection in severe COVID-19. To further understand the relevant functional status of each NK and T cell subsets between mild and severe patients, we conducted GEX analysis. We found T cell activation, migration, calcium ion signaling molecules were upregulated in CD8^+^ T cells from mild cases, but the virus responses, metabolites and energy generation, and translation initiation molecules was upregulated in CD8^+^ T cells from severe cases (Figure 3E and 3F). Thus, these data confirm our assumption that more effective CD8^+^ T cells in mild patients help to clear the viruses, while CD8^+^ T cells in severe patients are in preparation to proliferate.

### Characterization of single-cell TCR repertories in BALF from COVID-19 patients

When encountering antigens, specific B and T lymphocytes respond through clonal expansion to generate effector and memory cells with the same specificity to deal with the invaded pathogens. Expanded anti-viral T cell clones may exert protective or pathogenic functions during viral infections dependent on different situations. Here, we took advantages of single-cell TCR-seq to assess the status of clonal expansions in the BALF of patients. As expected, CCR7^+^ T cells as naive or central memory cells, showed little clonal expansion, while CD8^+^ effector T cells showed the highest expansion levels among different T cell subsets (Figure 4A and S5). Proliferating T cells and Treg cells also showed some expansion, but were less pervasive. Next, we examined the proportions of expanded clones between COVID-19 patients with mild and severe diseases. We found that the expansion levels in both total T and CD8^+^ T cells were significantly higher in mild disease group than the severe patients (Figure 4B and 4C, S5). In average, more than 50% CD8^+^ T cells in mild group were expanded clones, which likely represented the SARS-CoV-2-specific T cells. They also showed higher amplification index than those from severe patients (Figure 4C). At the individual levels, we also found the higher T cell clonality was consistently remained in the 3 mild patients as compared to the 3 severe patients (Figure 4D), supporting that highly expanded CD8^+^ T cell participated in resolving the SARS-CoV-2 infection. To confirm the functional status of expanded T cell clones, we performed GEX analysis between expanded CD8^+^ T cells versus non-expanded cells. We found increased expression of signaling and tissue resident genes including *XCL1, XCL2, ZNF683, HOPX, CXCR6* and *ITGAE*, etc., which further supports a superior effector functions of those expanded CD8^+^ T cells (Figure 4E).

**Figure 4.**
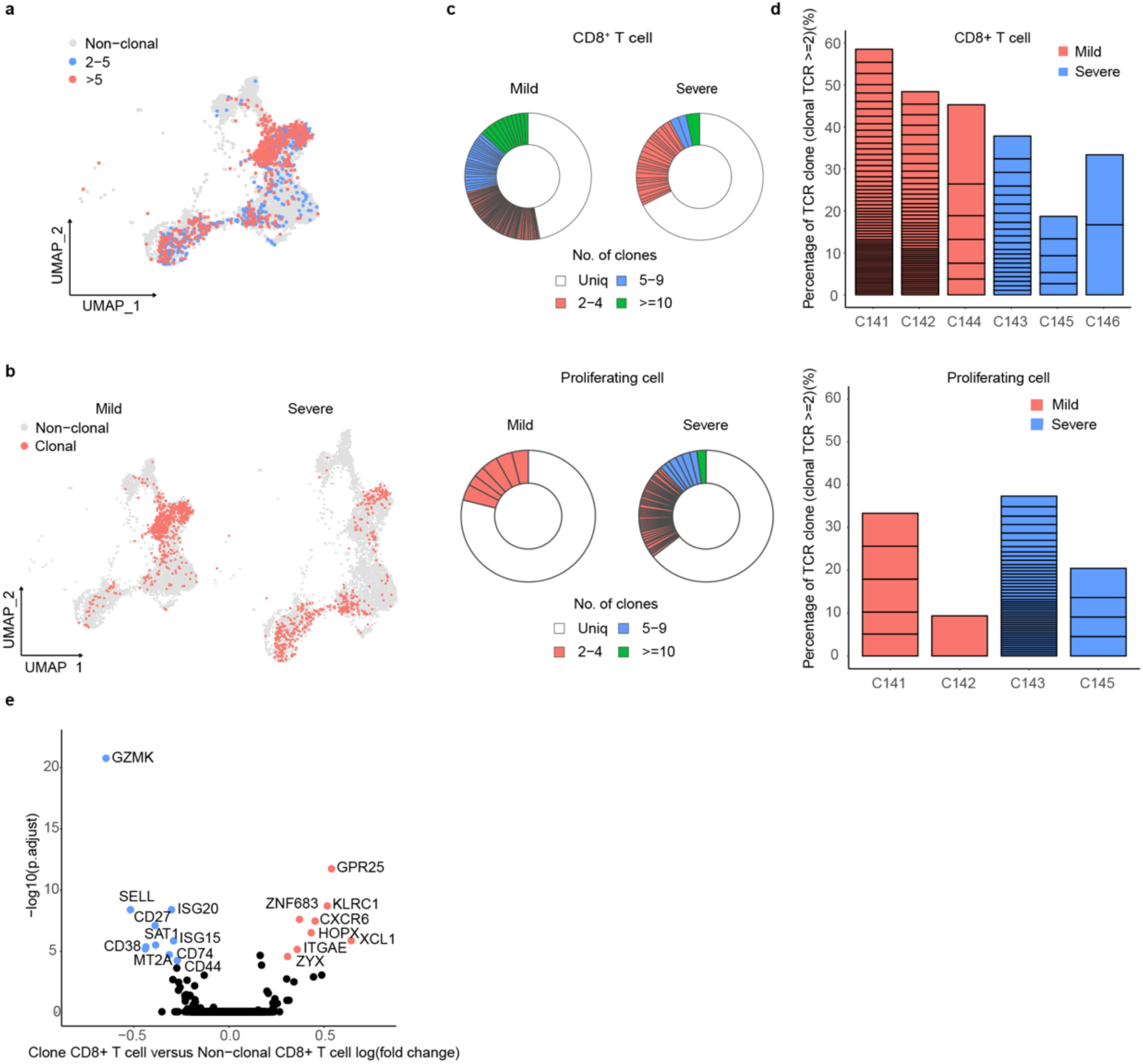
Analysis of the T cell clonal expansion in BALF of SARS-COV-2 infected patients. A. The UMAP plot shows the T cell expansion status in COVID-19 patients B. The UMAP projection of expanded clonal T cells between severe and mild patient group C. The pie charts show the clonal expansion status of CD8^+^ T and proliferating T cells in BALFs of severe and mild COVID-19 patients D. The bar plots show clonal expansion status of CD8^+^ T and proliferating T cells in BALFs from each individual sample E. The volcano plot shows the DEGs of expanded vs. non-expanded CD8^+^ T cells in BALFs of COVID-19 patients. A gene is considered significant with adjust p < 0.05. The top 8 up and down-regulated gene are labeled.

## Discussion

The clinical symptoms of COVID-19 varied from asymptomatic to ARDS, which has been similarly observed in SARS-CoV, MERS-CoV and influenza infections [7]. Host immune responses on some extent determine both protection and pathogenesis to the respiratory viral infections [11, 14]. A well co-ordinated innate and adaptive immune response may rapidly control of the virus, while a failed immune response leads to viral spreading, cytokine storm, and high mortality [15]. However, the immune players and the detailed molecular mechanisms related to these different processes are incompletely understood yet, in SARS-CoV-2 infection.

Previous studies have shown the significantly higher levels of inflammatory cytokines, chemokines and interferon stimulated genes associated with disease severity in SARS, MERS and COVID-19 patients [10, 16]. The increased infiltrating macrophages into the lung are identified in the autopsy and animal models, and are thought to be responsible for fueling the inflammation [17-19]. Macrophages are well known to contain heterogeneous subsets, including tissue resident and monocyte derived populations, covering a broad spectrum from M1 to M2 like phenotype [20]. However, little is known regarding the macrophage subsets and their molecular features in SARS-CoV-2-infected human lungs. Here, we identified 4 groups of lung macrophages which can be classified by FCN1, SPP1 and FABP4 markers according to recent reports [13]. Group 1 & 2 macrophages are FCN1^+^ and highly inflammatory. They express higher levels of interferon stimulated genes and multiple chemokines, *CCL2, CCL3, CCL5, IL-8, CXCL9, CXCL10 and CXCL11*, thus identifying the FCN1^+^ macrophages as the culprit for the deranged hyper-inflammation. We identified the group 3, SPP1^+^ macrophages likely represented a repaired but also a pro-fibrotic subset [12]. They may counteract the FCN1^+^ macrophages to dampen the inflammation. Their roles and associations with patient outcomes should be further investigated. The group 4 macrophages are FABP4^+^ AMs showing higher PPARγ activity which play important physiological roles in metabolizing lipid surfactant [20]. Here, we found that AMs consisted the principle lung macrophages in both controls and mildly infected patients, but was almost completely lost in severely infected lungs. The loss of AMs likely contributed to the failed lung functions. Together, these data improved our understanding of tissue macrophage heterogeneity in the immunopathogenesis of SARS-CoV-2 infection.

Adaptive immune system is specific and memorizes the pathogens, including two arms, the antibody and T cell responses. Inducing adaptive immunity is also the aim of vaccination. SARS studies showed that binding and neutralizing antibodies are elicited in SARS-CoV infections, but their association with clinical outcomes is unclear [21]. Robust antibody responses were developed in severely infected SARS patients [22]. However, it is debated whether antibody-dependent enhancement played roles in disease exacerbation [23, 24]. Memory T cell responses are induced and maintained in SARS-CoV infected subjects [25]. T cell immunity was also confirmed to be necessary for resolving the viral infection in mouse models [26, 27]. However, the roles of T cells, especially resident T cells, in controlling the disease severity of SARS-CoV infected patients are not proved yet. Here, we applied both scRNA-seq and TCR-seq to characterize the lung resident T cell populations. Our data not only revealed the presence of a larger population of CD8^+^ T effectors in mild group, but also showed that these CD8^+^ T cells contain highly expanded clones, indicating their SARS-CoV-2 specificity. Thus, we presented the fresh evidence that CD8^+^ T cell response likely holds the key for successful viral control in COVID-19 patients. This data will guide the future development of vaccine and potential T-cell immunotherapy for the severe viral respiratory infections.

The shortcomings of our study is that only 6 samples are able to be investigated and our endpoint study may not determine the triggering factors leading to different clinical outcomes. We propose that characterizing the earlier immune events and longitudinal analysis of immune dynamics of the infected patients may help gain more insights. These emerging high dimensional techniques, like scRNA-seq, mass cytometry and multiplex imaging tools [28, 29], will greatly improve human immunology studies using minimal precious tissue samples.

## Materials and Methods

### Patients

#### Ethics statement

This study was conducted according to the principles expressed in the Declaration of Helsinki. Ethical approval was obtained from the Research Ethics Committee of Shenzhen Third People’s Hospital. All participants provided written informed consent for sample collection and subsequent analyses.

#### Subjects and clinical sample collection

The six patients used in this study were recruited from the Shenzhen Third People’s Hospital in January, 2020. The demographic characteristics of these study populations are provided in Table 1. The patients included 3 mild cases and 3 severe cases, the median age is 49.5 years old, 5 male and 1 female. All patients had Wuhan exposure history and had cough and (or) fever as the first symptom. Diagnosis of SARS-CoV-2 was based on clinical symptoms, exposure history,chest radiography, and sputum, nasal swab and/or bronchoalveolar lavage fluid (BALF) viral PCR assays. Influenza A and B virus infection were excluded at the time of enrollment. The patient C142 has clinical symptoms, CT findings although SARS-CoV-2 PCR negative. She is a family member and close contact with C143.

### Quantitative reverse transcription polymerase chain reaction

The throat swab, sputum, nasal swab and BALF were collected from the respiratory tract of the patients at various time-points after hospitalization. Viral RNAs were extracted by Viral RNA was extracted from samples using the QIAamp RNA Viral Kit (Qiagen, Heiden, Germany). Then quantitative reverse transcription polymerase chain reaction (qRT-PCR) was performed using the primers and probes targeting ORF1ab and N genes of SARS-CoV-2 recommended by China CDC, and a commercial kit specific for SARS-CoV-2 detection (GeneoDX Co., Ltd., Shanghai, China). The specimens were considered tentatively positive if qRT-PCR Ct value lower than 37 is detected at any time-points during the hospitalization. Then the assay is repeated to confirm the positivity if the Ct value was similar or between 37∼40. If the repeat Ct was above 40, then the specimen was considered negative.

### Isolation of BALF cells

About 20 ml of BALF was obtained and placed on ice. BALF was processed within 2 hours and all operations were performed in BSL-3 laboratory. By passing BALF through a 100 µm nylon cell strainer to filter out lumps and then the supernatant was centrifuged and the cells were re-suspended in the cooled RPMI 1640 complete medium. Then the cells were counted in 0.4% trypan blued, centrifuged and re-suspended at the concentration of 2×10^6^ /ml for further use.

### Capturing, library construction and sequencing

Total 11 µl of single cell suspension and 40 µl barcoded Gel Beads were loaded to Chromium Chip A to generate single-cell gel bead-in-emulsion (GEM). The poly-adenylated transcripts were reverse-transcribed later. The single-cell capturing and downstream library constructions were performed using the Chromium Single Cell 5’ library preparation kit according to the manufacturer’s protocol (10x Genomics). Full-length cDNA along with cell-barcode identifiers were PCR-amplified and sequencing libraries were prepared and normalized to 3 nM. The constructed library was sequenced on BGI MGISEQ-2000 platform.

### Healthy Control Data

The lung scRNA-seq data from the healthy controls was acquired from the Gene Expression Omnibus (GEO) database under the series number GSE122960 [12], which contains data of lung tissue from eight lung transplant donors generated using 3’ V2 chemistry kit on Chromium Single cell controller (10xGenomics). Filtered feature-barcode matrix was used in the following analysis.

### ScRNA-seq data alignment and sample aggregating

The Cell Ranger Software Suite (Version 3.1.0) was used to perform sample de-multiplexing, barcode processing and single-cell 5’ UMI counting with human GRCh38 as the reference genome. Specifically, splicing-aware aligner STAR [30] was used in FASTQs alignment. Cell barcodes were then determined based on distribution of UMI count automatically. Finally, gene-barcode matrix of all 6 donors and 8 healthy control was integrated with Seurat v3 [31] to remove batch effect across different donors. Following criteria were then applied to each cell, i.e., gene number between 200 and 6000, UMI count above 1000 and mitochondrial gene percentage below 0.1. After filtering, a total of 81447 cells (5436/3858/5235/5191/5341/4300/7516/6750 cells for healthy control; 3542/3411/363 cells for mild patients; 17340/11872/1292 cells for severe patients) were left for following analysis.

### Dimensionality reduction and clustering

The filtered gene-barcode matrix was normalized with LogNormalize methods in Seurat and analyzed by principal component analysis (PCA) using the top 2, 000 most variable genes. Then Uniform Manifold Approximation and Projection (UMAP) was performed on the top 50 principal components for visualizing the cells. Meanwhile, graph-based clustering was performed on the PCA-reduced data for clustering analysis with Seurat v3.

### Differential analysis for clusters

MAST [32] in Seurat v3 was used to perform differential analysis. For each cluster, differentially-expressed genes (DEGs) were generated relative to all of the other cells.

### Single cell trajectory analysis

Slingshot [33] was used to perform pseudotime inference for the four myeloid cell groups.

### Regulatory network inference

Single cell regulatory network for 4 myeloid groups was constructed with SCENIC [34]. Specifically, GRNBoost2 (https://github.com/tmoerman/arboreto) in pySCENIC was applied to infer gene regulatory networks from expression data. The regulators-group heatmap was generated with R package AUCell.

### Gene functional annotation

For DEGs, Gene ontology (GO), KEGG pathway analyses and Gene Set Enrichment Analysis (GSEA) [35] were performed with clusterProfiler [36], which supports statistical analysis and visualization of functional profiles for genes and gene clusters. In GSEA analysis, 50 hallmark gene sets in MSigDB [37] were used for annotation.

### TCR V(D)J sequencing and analysis

Full-length TCR V(D)J segments were enriched from amplified cDNA from 5’ libraries via PCR amplification using a Chromium Single-Cell V(D)J Enrichment kit according to the manufacturer’s protocol (10x Genomics). The TCR sequences for each single T cell were assembled by Cell Ranger vdj pipeline (v3.1.0), leading to the identification of CDR3 sequence and the rearranged TCR gene. Cells with both TCR alpha and beta chains were kept and cells with only one TCR chain were discarded. The quality of TCR-seq was listed in Table S2. Then the clonality of the different T cells defined by scRNA-seq data was explored.

## Data Availability

Raw data will be available in GEO.

